# Physician Perceptions of Catching COVID-19

**DOI:** 10.1101/2021.01.15.20249089

**Authors:** P. Murali Doraiswamy, Mohan Chilukuri, Dan Ariely, Alexandra R. Linares

## Abstract

**Background:** Risk perception, influenced and biased by multiple factors, can affect behavior.

**Objective:** To assess the variability of physician perceptions of catching COVID-19.

**Design:** Cross sectional, random stratified sample of physicians registered with Sermo, a global networking platform open to verified and licensed physicians.

**Main outcome measures:** The survey asked: “What is your likelihood of catching COVID-19 in the next three months?*”* The physicians were asked to give their best estimate as an exact percentage.

**Results:** The survey was completed by 1004 physicians (40 countries, 67 specialties, 49% frontline [e.g. ER, infectious disease, internal medicine]) with a mean (SD) age of 49.14 (12) years. Mean (SD) self-risk estimate was 32.3% ± 26% with a range from 0% to 100% (Figure 1a). Risk estimates were higher in younger (<50 years) doctors and in non-US doctors versus their older and US counterparts (p<0.05 for all) (Figure 1b). Risk estimates were higher among front line versus non-frontline doctors (p<0.05). Risk estimates were higher for women than men (p<0.05) among respondents (60%) reporting gender.

**Conclusions:** To our knowledge, this is the first global study to document physician risk perceptions for catching COVID-19 and how it is impacted by age, gender, practice specialty and geography. Accurate calibration of risk perception is vital since both over- and underestimation of risk could impact physician behavior and have implications for public health.

## Introduction

Despite more than 87 million cases of COVID-19 worldwide, the number of infected physicians is not fully known. While efforts are underway to chart the actual infection risk among physicians (1-7), it is equally important to understand perceived risk which influences behavior. Risk perception is a subjective process in which people use heuristics (shortcuts) to evaluate information (8). Such heuristics are influenced by many factors including personal experiences and beliefs – one reason why people see the same risk differently. This process is subject to cognitive biases – for example, people often overestimate risks that are beyond their control and underestimate risks that they undertake willingly (8,9).

## Methods

This is an analysis of an anonymous, cross-sectional, random, stratified, IRB exempt survey, done September 9-15, 2020, of verified physicians registered with Sermo, a digital platform for medical crowdsourcing. Following online informed consent, this survey aimed to sample 1,000 doctors equally divided between USA, EU and rest of the world (RoW) on many topics of which one question pertained to perceived risk. The self-risk question was *“What is your likelihood of catching COVID-19 in the next three months?”* The physicians were asked to give their best estimate as an exact percentage. Data were analyzed with JMP Pro15 and Protobi.

## Results

The survey was completed by 1004 physicians (40 countries, 67 specialties, 49% frontline [e.g. ER, infectious disease, internal medicine]) with a mean (SD) age of 49.14 (12) years. Mean (SD) self-risk estimate was 32.3% + 26% with a range from 0% to 100% (Figure 1a). Risk estimates were higher in younger (<50 years) doctors and in non-US doctors versus their older and US counterparts (p<0.05 for all) (Figure 1b). Risk estimates were higher among front line versus non-frontline doctors (p<0.05). Risk estimates were higher for women than men (p<0.05) among respondents (60%) reporting gender.

**Figure 1A.**
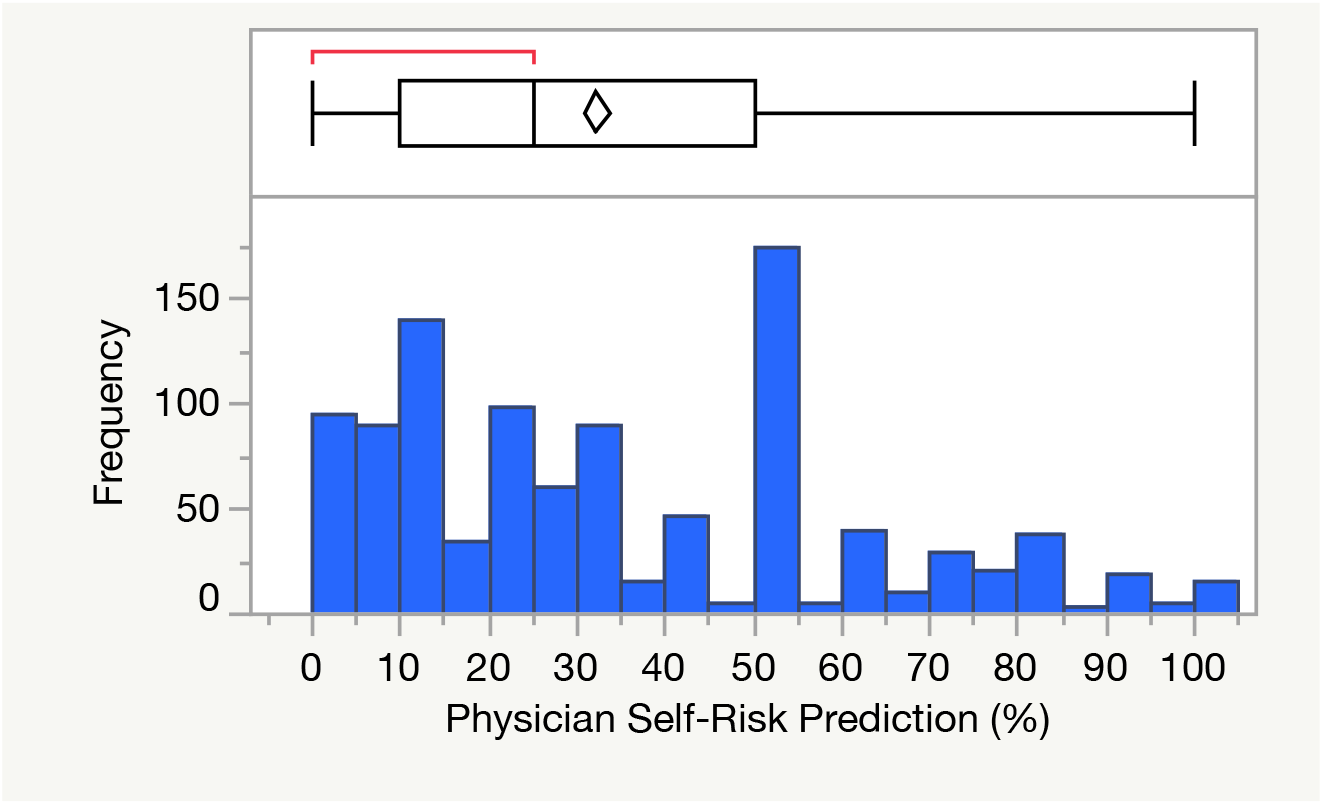
Distribution of risk prediction for overall sample (N=1004). Upper panel is a line box whisker and bottom panel shows the frequency distribution.

**Figure1B.**
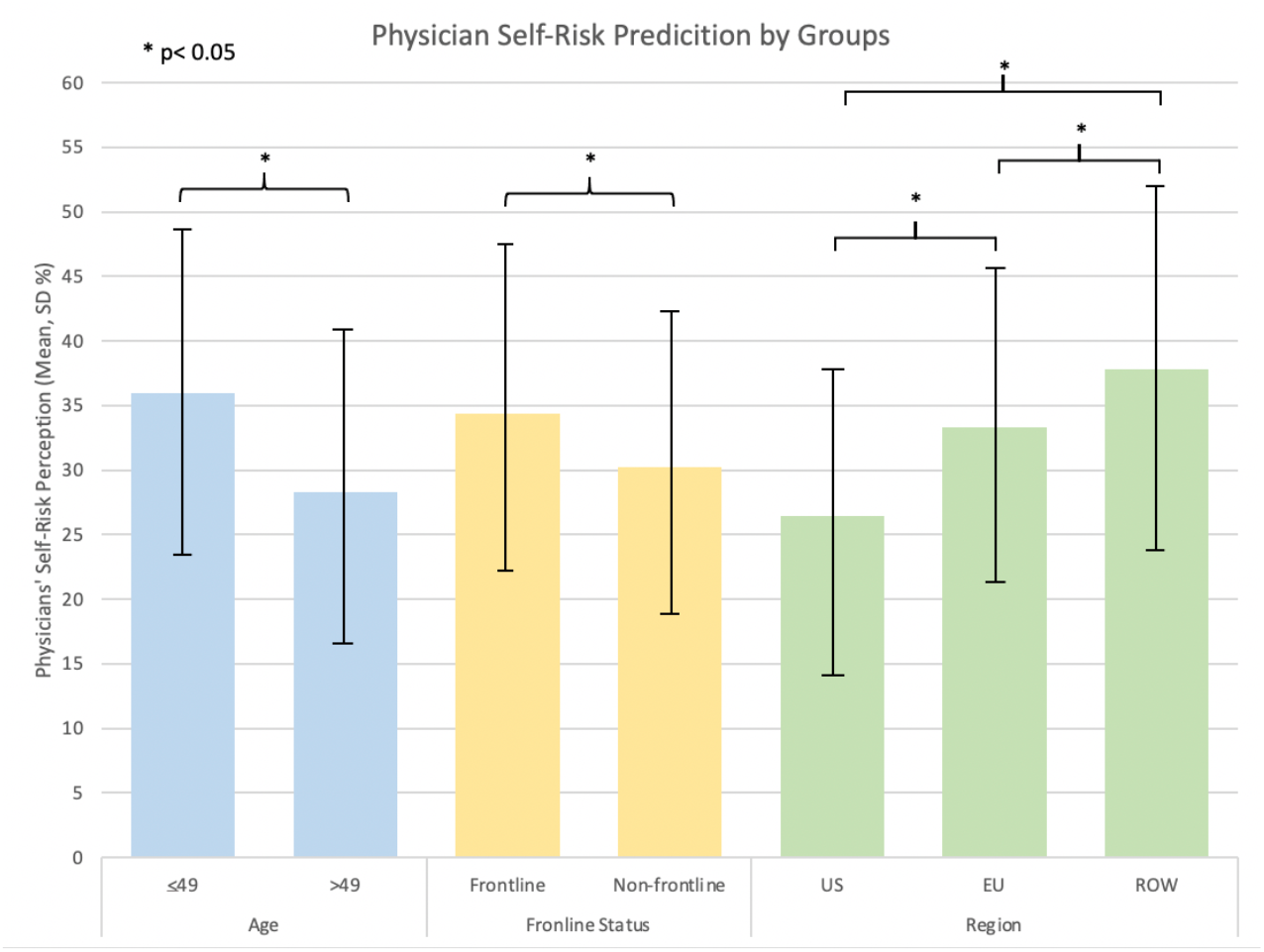
Mean (SD) risk estimates by age, frontline status, and geographic region.

## Discussion

The average 3-month risk perception estimate of 32%, if true, would mean that one-third of all physicians worldwide (including some 258,000 doctors in the US) will be infected by December 2020. This is higher than the reported physician SARS-CoV-2 seroprevalence rates of 3.6-19% (1-7). This survey also finds a high variability of risk perception and differences by age, gender and geography – suggesting factors other than actual risk may be contributing.

Measuring and calibrating risk perception correctly is important since there are consequences to both overestimation and underestimation (8,9). Overestimation of risk can lead to greater compliance with safety regulations but also anxiety and avoidance of necessary activities (e.g., routine physical exams). Underestimation can raise a physician’s risk for catching COVID-19 and/or becoming super-spreaders. Physician risk perceptions may also influence how they educate patients and formulate public health safety policies. Lastly, this survey provides insights into how the broader health worker community may perceive infection risk.

Despite limitations such as cross-sectional nature, single question, and inability to control for all possible confounders, this survey provides foundational insights into physicians’ risk perception for COVID-19. Prospective studies of both actual and perceived risk across different medical specialties may enhance occupational safety.

## Data Availability

Data will be made available upon request.

## Ethics Declaration

This anonymous survey was conducted in September 2020 following an online informed consent process. This was a broad survey across many topics of which one question pertained to immunology. De-identified data was analyzed for this report. This was reviewed and deemed exempt research by the Duke University Medical Center Institutional Review Board.

## Acknowledgements

The authors thank Sermo and the physicians who took part in the survey.

